# Spatial determination of COVID-19 mortality

**DOI:** 10.1101/2022.09.02.22279526

**Authors:** GC Arun

## Abstract

COVID-19 has affected at the global scale. However, its impacts are not evenly distributed. The article aims to explore the spatial determination of the COVID-19 related death. The data for the analysis has been accessed from the World Health Organization (WHO). Both descriptive and statistical analysis has been done to assess the COVID-19 related death and spatial explanation. The regression models suggested the explanatory power of spatial difference in the COVID-19 related death. However, further addition of various COVID-19 vaccine did not produce expected result.

## Introduction

SARS-CoV-2 which is now widely known as COVID-19 virus was reported by the Wuhan Municipal Health Commission of China on 31 December 2019. Later, on 11 March 2020, the World Health Organization (WHO) declared the viral disease as a pandemic. Till now, millions of cases were confirmed from around the world. However, distribution of new cases and COVID-19 related cases are disproportional. There are several findings suggested there are determinants of COVID-19 related deaths. Cancer diseases and COVID-19 related deaths were found correlated[1, 2]. Another study identified that the level of the economic growth can determine the COVID-19 related death rate[3]. COVID-19 is also considered as the most individualist and has disproportional impact on gender and age[4]. Besides age and gender, ethnicity is also found important determinant for COVID-19 mortality[5]. So, it is clear that there are several determinants of COVID-19 mortality. The question is whether COVID-19 related mortality has any spatial explanation. Therefore, the study attempts to analyze the regional differences of COVID-19 mortality.

## Materials and Methods

To assess the spatial determination of COVID-19 related mortality, both descriptive and the statistical tools were employed. Linear regression model was used for econometric analysis. The data for this study was accessed from the World Health Organization (WHO) data base (https://covid19.who.int/data) on July 27, 2022. The dataset contains country’s name, WHO region, cumulative total cases, cumulative total cases per 100,000 population, newly reported cases in last 7 days, newly reported cases in last 7 days per 100,000 population, newly reported cases in last 24 hours, cumulative total death, cumulative total death per 100,000 population, newly reported death in last 7 days, newly reported death in last 7 days per 100,000 population, and newly reported death in last 24 hours. The original dataset contains 238 rows for spatial units including global aggregation.

## Results and Discussion

Data exploration is very important part for the analysis. For this study, global total and rows containing zero cumulative covid19 cases were discarded. From this process, a total 233 countries (entries) were considered. Figure 1 presents the distribution of countries in the given regions.

**Figure 1:**
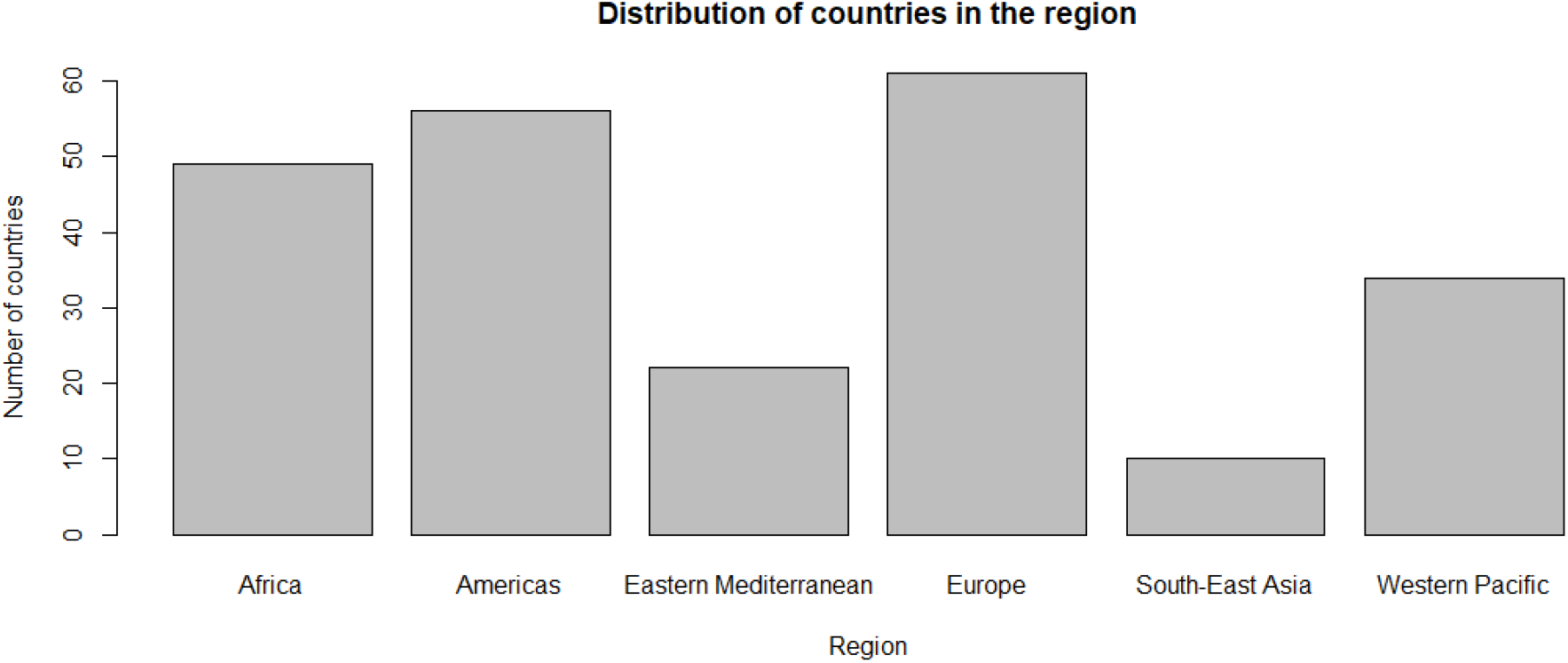
Distribution of countries within the given regions

From the available variables, the following variables (Table 1) were considered for the study. For the simplicity, the regions were considered as they are categorized by the WHO. They are Africa, Americas, Eastern Mediterranean, Europe, South-East Asia and West Pacific. It is different from the conventional regionalization under the content basis. The COVID-19 cases and mortality were standardized with per 100,000 populations. Table 1 describes the variables. There are maximum numbers of countries in the Europe region and minimum in the South-East Asia.

**Table 1:** Description of variables

| Region | Number of Countries | Covid19 Cases Per 100,000 |  | Covid19 death per 100,000 |  |
| --- | --- | --- | --- | --- | --- |
| Africa | 49 | Minimum | 37.61 | Minimum | 0 |
| Americas | 56 | 1st Quarter | 1780.12 | 1st Quarter | 15.07 |
| Eastern Mediterranean | 22 | Median | 11763.63 | Median | 81.51 |
| Europe | 61 | Mean | 17578.57 | Mean | 117.69 |
| South-East Asia | 10 | 3rd Quarter | 29087.6 | 3rd Quarter | 186.95 |
| Western Pacific | 34 | Maximum | 70926.02 | Maximum | 649.12 |

The maximum cumulative covid-19 cases per 100,000 was observed in Faroe Islands under Europe region. Similarly, Europe is found the region which contains maximum number of countries having higher incidence of covid-19 cases. European region has 43 countries which have above the mean covid-19 cases per 100,000 population. In total, 89 countries were reported to have more than the mean covid-19 cumulative cases per 100,000 population. Figure 2 presents the number of countries in different regions where cumulative covid-19 cases per 100,000 is more than global mean.

**Figure 2:**
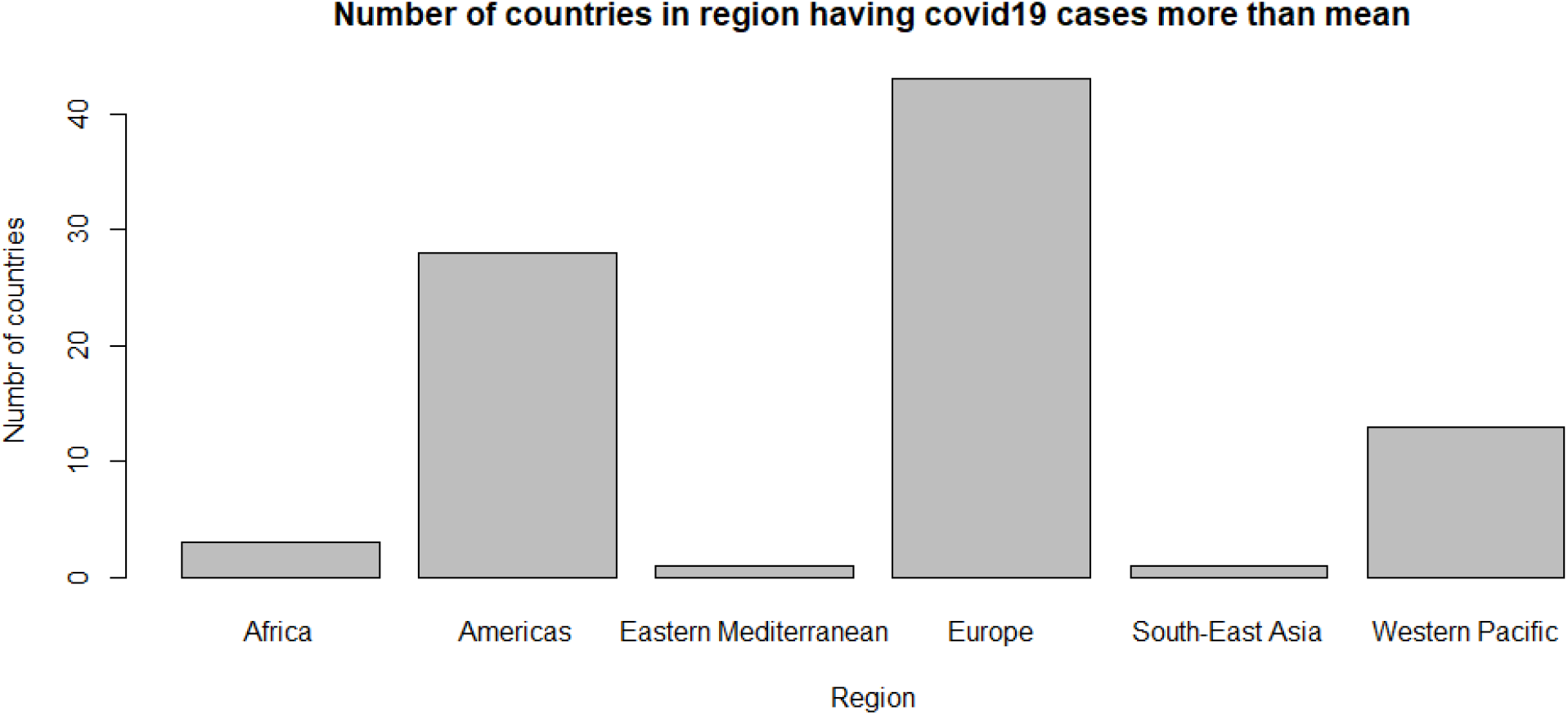
Number of countries having more than global mean cumulative covid19 cases per 100,000

Similarly, the least cumulative covid-19 cases per 100,000 was observed in Niger under African region. Total 143 countries have less than global mean cumulative covid-19 cases per 100,000 population. African region has majority of countries having less than global mean cumulative cases of covid-19 per 100,000 population whereas Europe has just 18 such countries. Figure 3 depicts the details.

**Figure 3:**
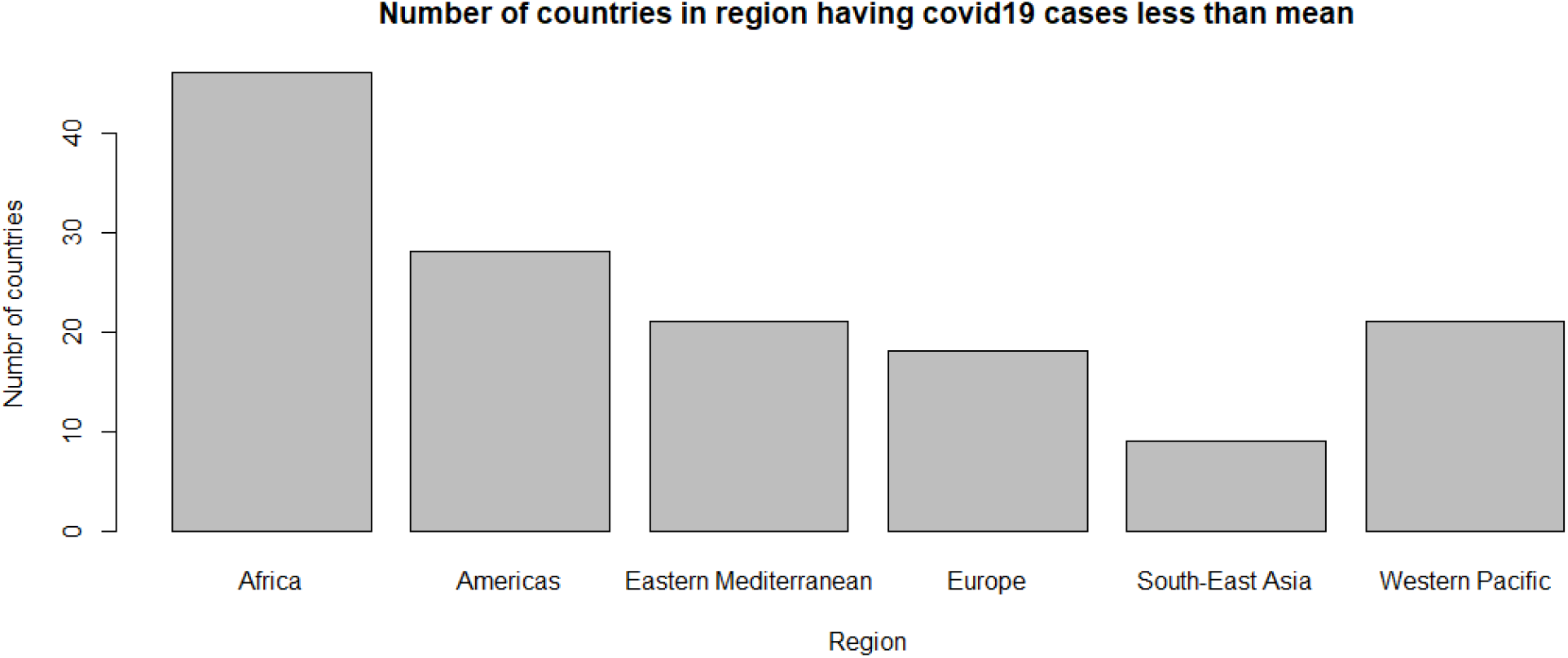
Number of countries having less than global cumulative covid19 cases per 100,000 population

Figure 4 presents the distribution of cumulative covid-19 cases in the different regions. The figure suggests that Europe has higher mean as well as higher range as compared to other regions.

**Figure 4:**
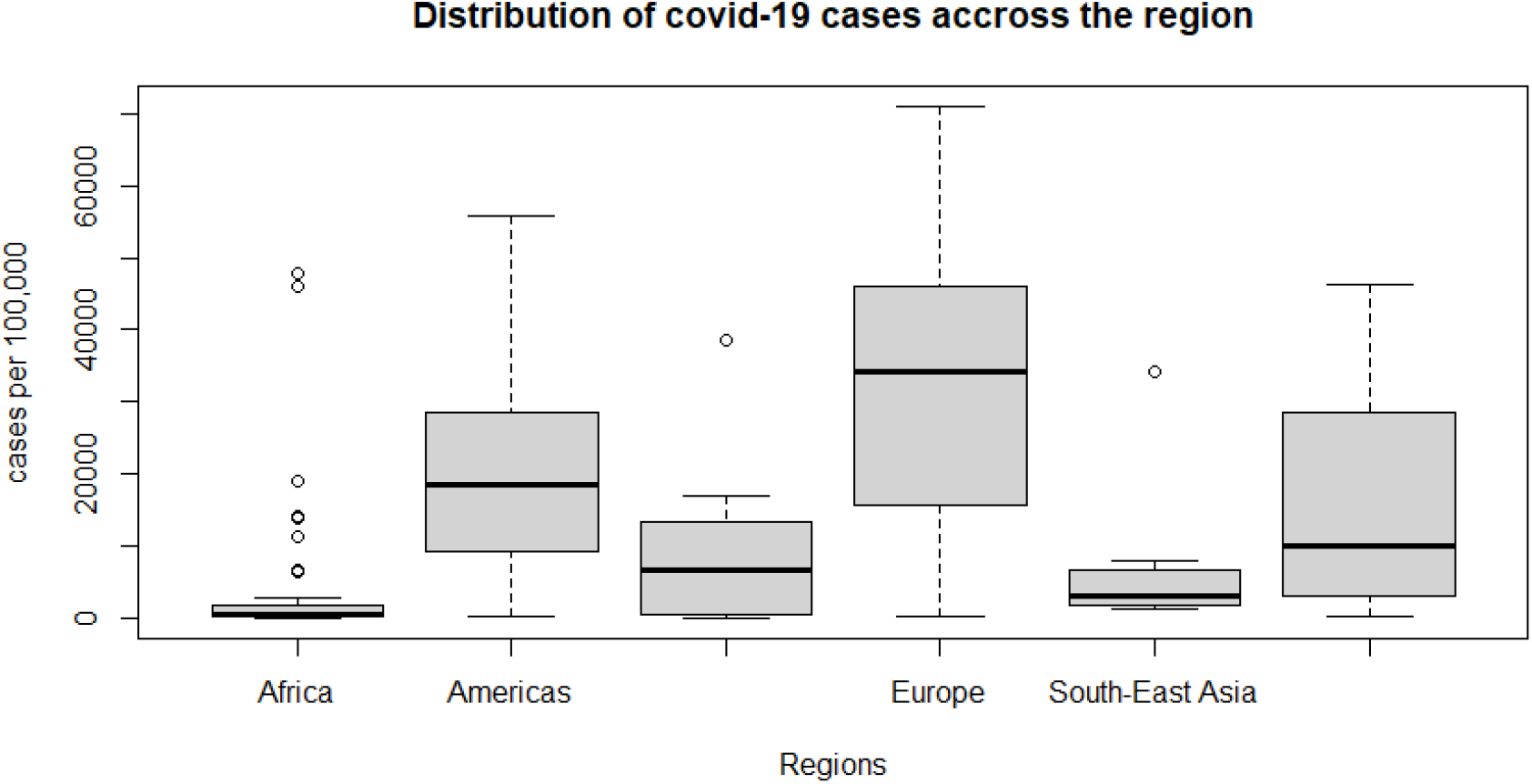
Distribution of cumulative covid19 cases around the regions

Peru observed maximum covid-19 related death per 100,000 population and Americas as a region. Total 91 countries observed number of deaths above the global mean per 100,000 population. Total 35 countries in Americas have covid-19 death more than the global mean per 100,000 population. Interestingly, none of the countries in the South-East Asia region has covid-19 related death more than the global average. Figure 5 describes the distribution of covid-19 related death among different regions.

**Figure 5:**
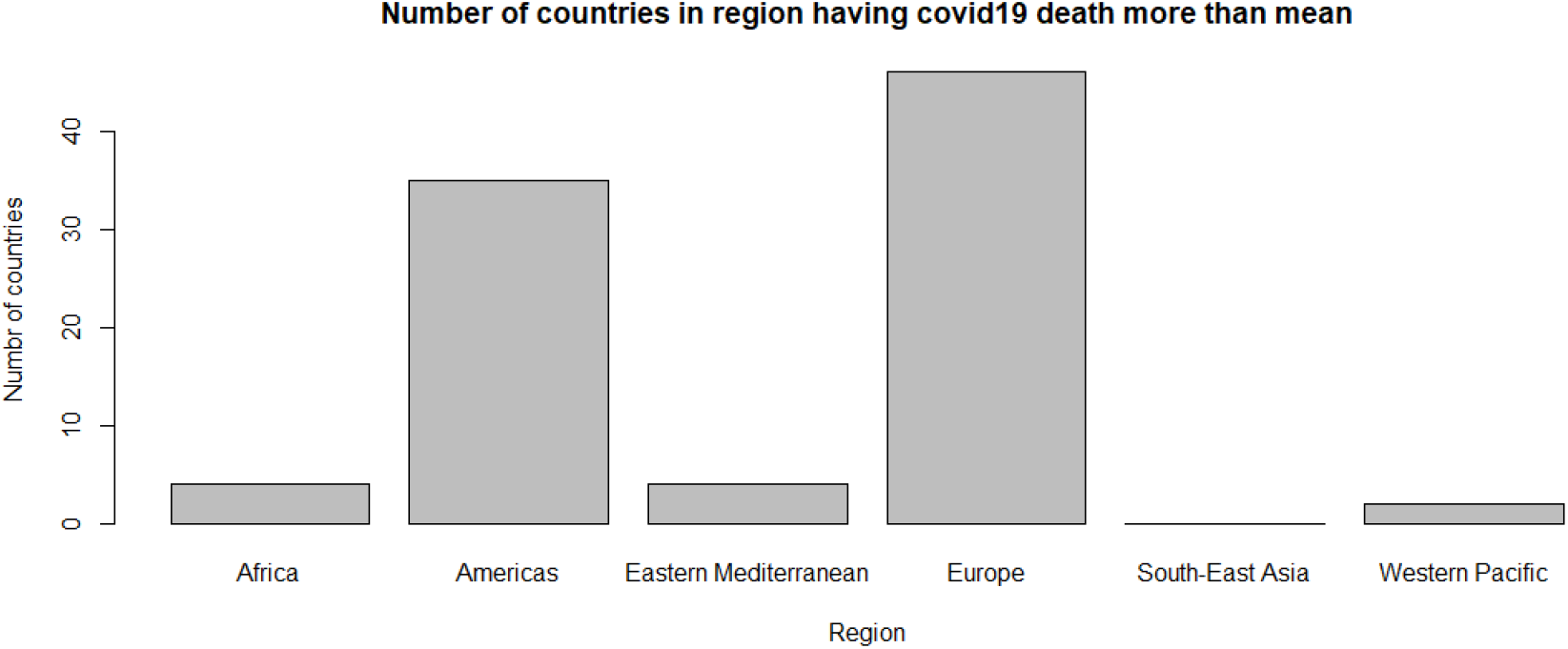
Distribution of countries in terms of covid19 related death in per 100,000 population

Falkland Islands (Malvinas), Holy See, Marshall Islands, Niue, Pitcairn Islands, and Tuvalu do not have any covid-19 related death. Total 91 countries have covid-19 related death more than the global average per 100,000 population. Europe has total 46 countries where covid-19 related death is more than the global mean. Figure 6 presents the distribution of countries in different regions where covid-19 related death is less than the global mean.

**Figure 6:**
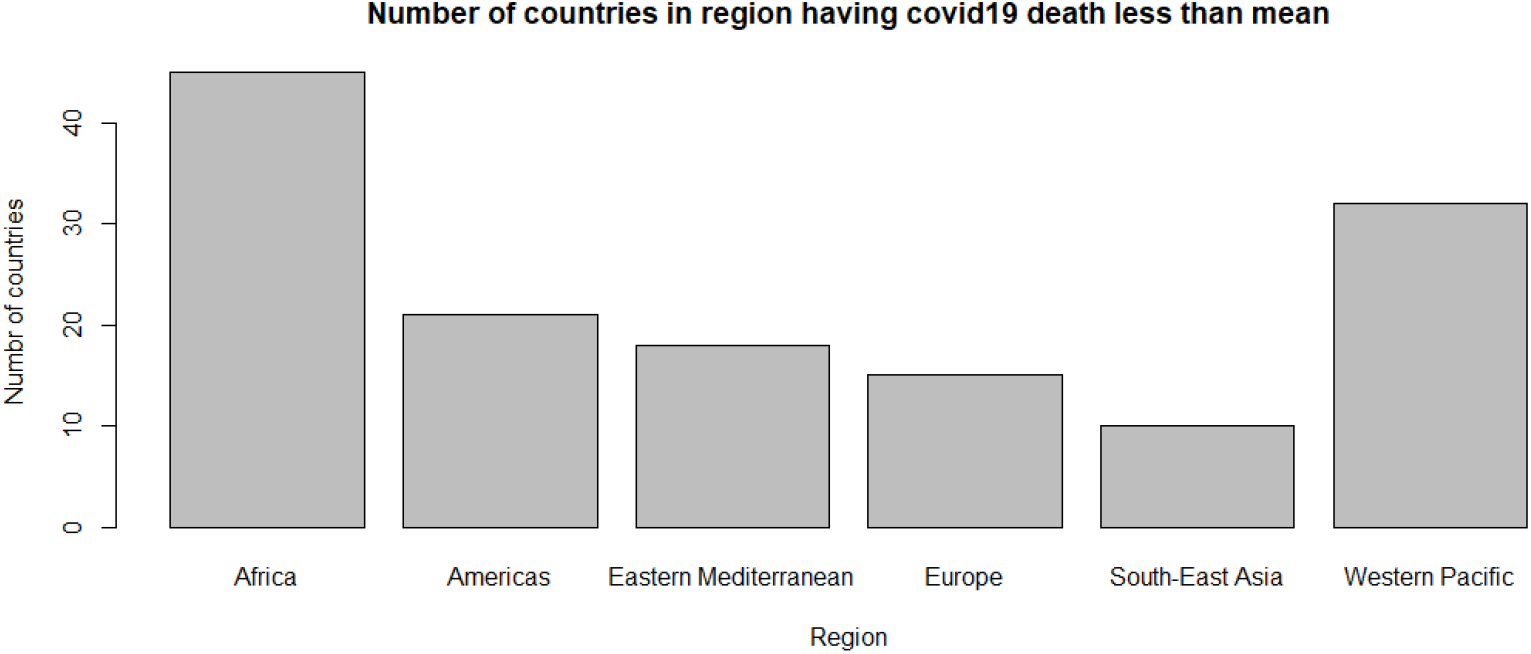
Regions having less than mean covid19 related death

The Figure 7 presents the distribution of covid-19 related death across the different regions. The figure suggests that Europe has higher mean and range in terms of covid-19 related death per 100,000 population.

**Figure 7:**
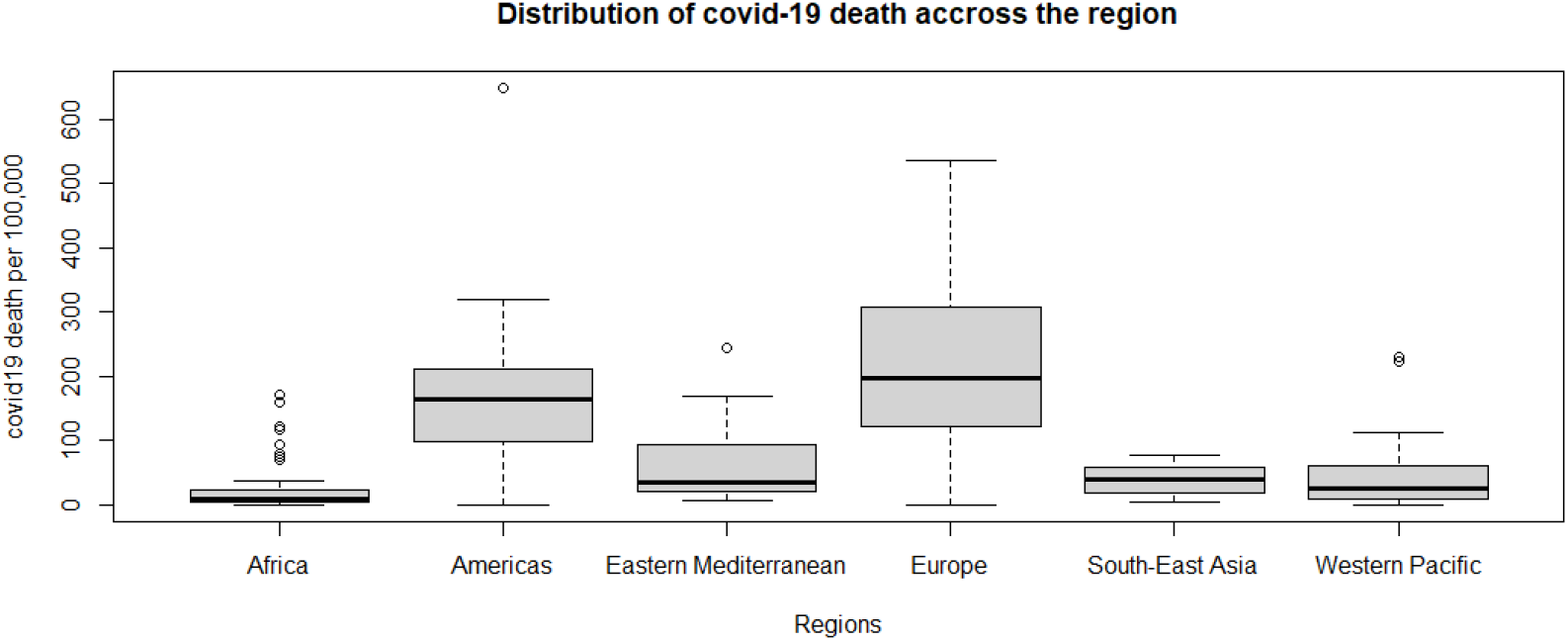
Distribution of covid19 related death across the region

### Regression Model

The ordinary least square method has been employed for econometric analysis. Total 232 observations have been included for the analysis. The cumulative covid-19 related death per 100,000 population was considered as a regressend and the cumulative covid-19 cases per 100,000 population were taken as regressor. The model is found highly significant with F-statistics (6, 225) 28.037. The R squared value was found 0.428 which means the model can explain 42.8 percent variations in the covid-19 related death per 100,000 population jointly by region and cumulative covid-19 case per 100,000 population. In case of region, Africa region is considered as a reference region. The regression analysis suggests that being in Americas region is 122.937 times more likely to encounter with covid-19 death as compare to being in Africa and is statistically significant too. Likewise, being in Europe, encounter with covid-19 related death will increase by 165.952 times as compared to being in Africa.

Regional differences in the covid-19 related death are an interesting phenomenon. The differences might have raised due to several reasons. Those regions have genetic variation[6-11]. An interesting finding was found in a study within the USA. The study suggested that mortality in urban region of the USA is higher than that of rural area[12]. Another study has suggested that air quality, demographics, global interconnectedness, urbanization trends, historic trends in health expenditure and policy measure for covid-19 mitigation have influence on mortality difference in EU countries[13]. Some studies have identified the similarity in the mortality within the reason[14]. Another study identified that GDP per capita, share of older people in the population and unemployment are related with COVID-19 death in EU region[15].

Some studies suggested that lower COVID-19 mortality rate in Africa is contributed by lower population mean age[16], lower life expectancy, lower pre-COVID-19 era 65 years and more mortality rate and smaller pool of people surviving and living with cardiovascular disease[17].

To understand further on the spatial difference on the covid-19 related death, covid-19 vaccination was also considered. Various models were developed and among them seven are presented in Table 3.

**Table 2:**
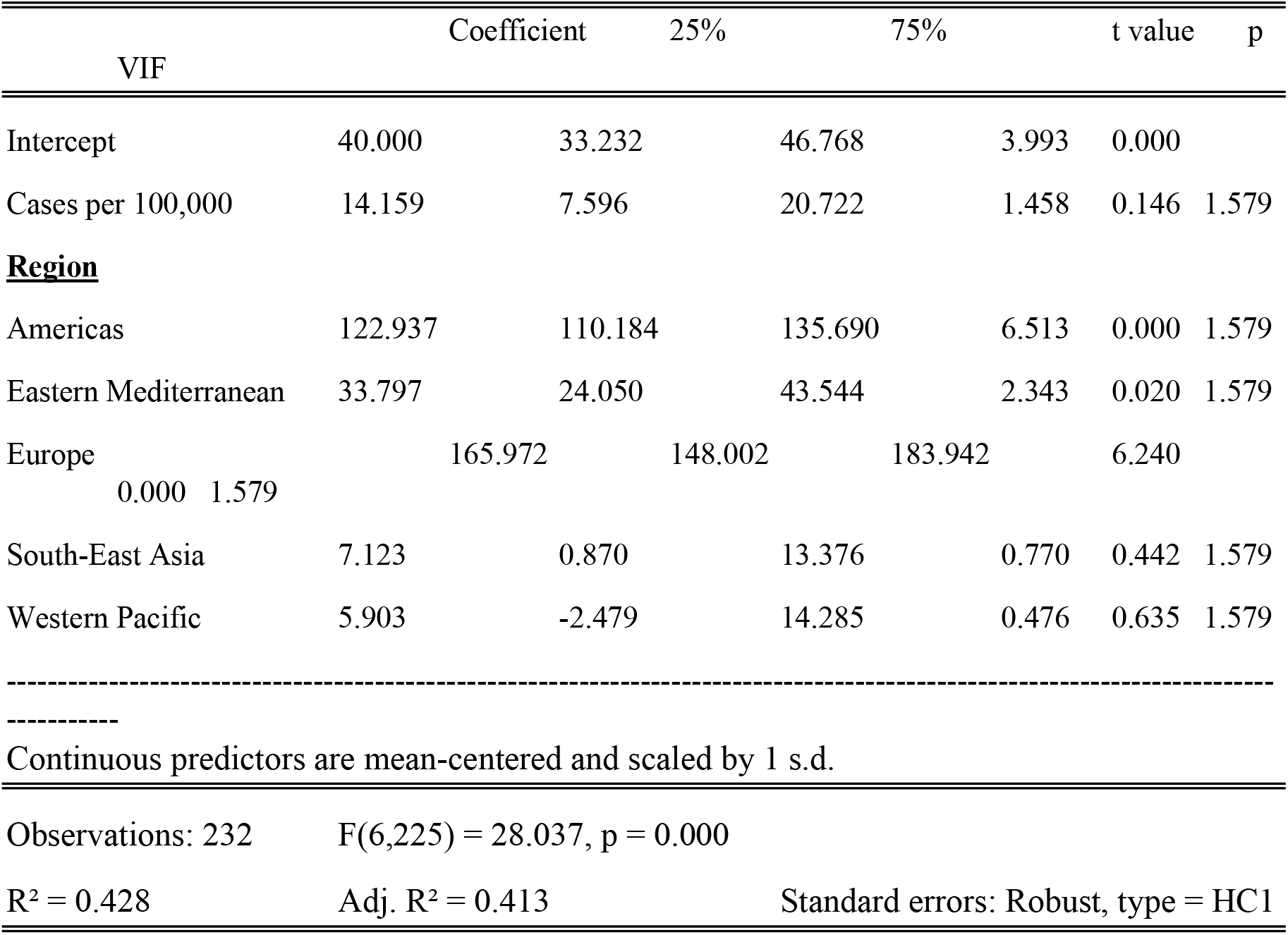
OLS model result

**Table 3:**
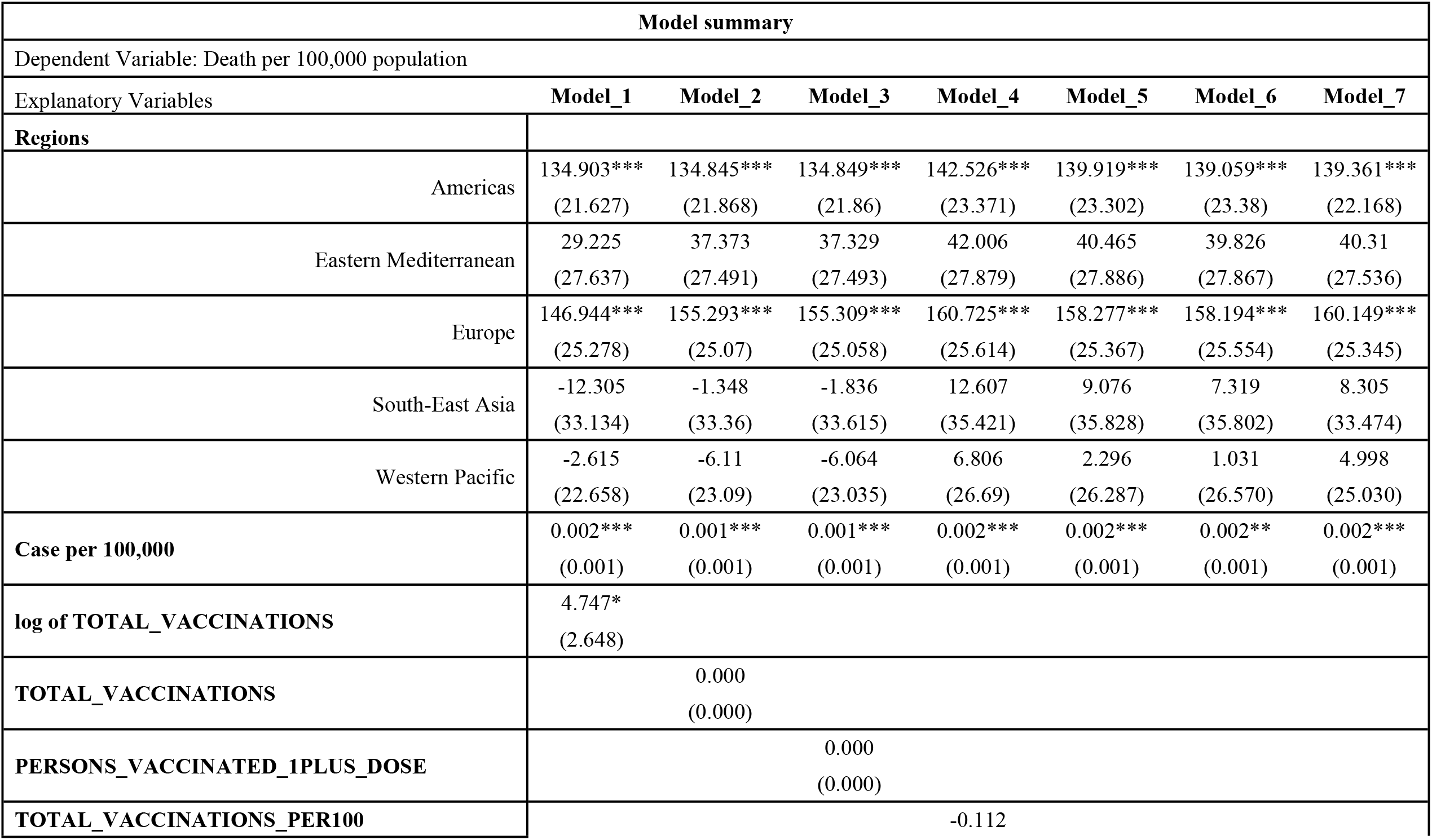
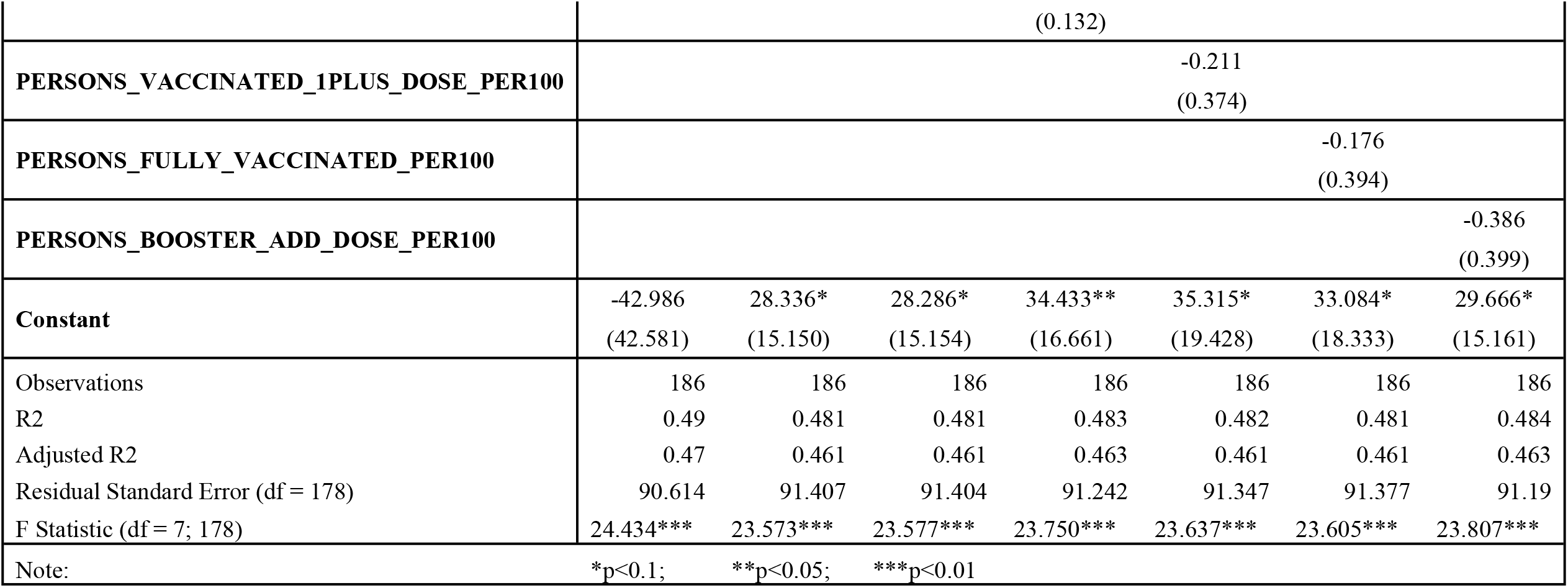
Model Summary of spatial variation with vaccination

In the base model presented before, following variables has been added to the model to compare for the better result.

a. Log of total vaccinations
b. Total vaccinations
c. Persons vaccinated with more than one doses
d. Total vaccinations per 100
e. Persons vaccinated with more than one doses per 100
f. Persons fully vaccinated per 100
g. Persons booster and additional doses per 100

All models are found significant at 5 percent level. However, value of R square is different. The Model_1 (with log of total vaccination) has the highest R square value and the lowest residual standard error. Further, AIC criteria was also employed. Table 4 presents the value of AIC. From the AIC criteria also Model 1 was found out performing other models. However, in Model 1, log of total vaccination has produced positive coefficient. Although, the coefficient is not significant, Model 4, Model 5, Model 6 and Model 7 have negative coefficient for the vaccination data.

**Table 4:** AIC of various models

| **Models** | **Degree of Freedom** | **AIC** |
| --- | --- | --- |
| Model 1 | 9 | 2214.127 |
| Model 2 | 9 | 2217.367 |
| Model 3 | 9 | 2217.355 |
| Model 4 | 9 | 2216.697 |
| Model 5 | 9 | 2217.125 |
| Model 6 | 9 | 2217.247 |
| Model 7 | 9 | 2216.482 |

Poor prediction of vaccination data on the mortality might have caused due to uneven distribution of covid-19 vaccine even within the region. A separate in-depth analysis is necessary for analysis of vaccination data and covid-19 related death, considering its sensitivity.

## Conclusion

Covid-19 related death has various determinants at the micro-level. However, at the general level, the spatial distribution might have some explanatory power. Therefore, this study has been carried out to analyze whether the covid-19 related death has any spatial explanation. The regression analysis, shows that the regions have significant explanatory power. In fact, region or spatial unit has aggregate information on various determinants of covid-19 related deaths. Further, various vaccination data has been added in the model. However, the explanatory power of the vaccination data in the given dataset was not found strong enough. Only one model out of seven yield the significant result but with positive sign on the coefficient. However, other models yielded negative coefficient but with no significant results. This article presents the overall distribution of covid-19 related death. It enables global policy makers to provide bird-eyes views for further interventions.

## Data Availability

XXX

WHO

## Notes

### Competing Interest Statement

The authors have declared no competing interest.

### Clinical Trial

NA

### Clinical Protocols

NA

